# Asymptomatic *P. falciparum* Infection is Not Associated with Exposure to Soil Transmitted Helminths in Children from a Multi School-Based Study in Esse, Cameroon

**DOI:** 10.1101/2025.09.18.25336099

**Authors:** Lauren Lajos, Balotin Fogang, Anne Jensen, Derrick Atchombat, Douglas H. Cornwall, Christiane Donkeu, Chris-Marco Nana-Mbianda, Celine Slam, Hugues Clotaire Nana Djeunga, Bin Zhan, Paul Olivier Koki Ndombo, Lawrence S. Ayong, Tracey J. Lamb

## Abstract

Asymptomatic carriage of *Plasmodium falciparum* is a major public health threat hindering malaria eradication. Many areas with ongoing malaria transmission are co-endemic for soil transmitted helminths (STH). Proteins secreted by helminths can regulate host inflammatory immune responses as a survival strategy. Given that malaria is a disease mediated by inflammation, we tested the hypothesis that STH infection and/or exposure might be associated with an asymptomatic phenotype of *Plasmodium* infection. We performed a one-month longitudinal study of 134 primary school children across 3 school-based study sites in Esse, Centre Region, Cameroon. At our initial screening time point, 94.8% of children were microscopy positive for *P. falciparum* infection and 85.8% had asymptomatic microscopic *P. falciparum* infection. A total of 87.4% of children had serologic positivity for at least one STH recombinant antigen. Comparing children with asymptomatic malaria and uncomplicated symptomatic malaria at baseline, we found no significant difference in the percentage of children with STH exposure (85.7% vs 90.9%, p >0.05). Daily temperature checks were performed over the course of one month to assess whether children with asymptomatic malaria developed uncomplicated symptomatic malaria. Of the children that developed uncomplicated malaria, development of fever was associated with increased reactivity to STH antigens. No correlation was found between anti-STH antibody level and *P. falciparum* load (p > 0.05), and no association was observed between STH exposure and persistent asymptomatic *P. falciparum* infection. This data suggests that STH exposure is not a major factor that contributes to the asymptomatic carriage of *P. falciparum* in children.

## INTRODUCTION

An estimated 1.5 billion people worldwide are infected with soil transmitted helminths (STH), including over 900 million children living in areas that are endemic for malaria^1^. In 2023 there were approximately 600,000 deaths from malaria, the majority of which occurred in children under 5 years old^2^. These diseases persist as major public health threats in part because most infected individuals in endemic regions are asymptomatic, allowing for uncontrollable disease transmission due to lack of early detection and treatment^3, 4^.

Although increasingly described as a widely prevalent and important public health threat, a standard definition for “asymptomatic malaria” is lacking^5^. However, this term is generally accepted to indicate *Plasmodium spp.* parasitemia of any density (microscopic or submicroscopic) in the absence of fever or other overt clinical symptoms in individuals who have not received recent anti-malarial treatment^5^. Malaria is a disease which is predominantly caused by inflammatory responses to *Plasmodium* parasites^6^. The mechanisms that underpin the asymptomatic carriage of the *Plasmodium* parasites are poorly understood but likely lead to down-regulation of some components of the inflammatory response in the face of ongoing *Plasmodium* infection.

Without treatment, certain STH can cause chronic infections lasting from several years to decades^7, 8, 9^. Depending on species, STH induce potent immunoregulatory immune responses that dampen the immune mechanisms that lead to expulsion of the adult helminths from the gastrointestinal tract^10^. This is achieved by several evolutionary adaptions including the secretion of homologs of immunoregulatory cytokines such as transforming growth factor-β (TGF-β)^11, 12, 13, 14^. A meta-analysis of *P. falciparum* infected individuals^15^ has shown that STH co-infections may impact control of *Plasmodium* parasites and the development of malarial anemia, but this does not occur in all cases^16^. Although it has been suggested that individuals with STH infections may be more likely to have asymptomatic / uncomplicated malaria^15^ it is not clear the extent to which asymptomatic carriage of *Plasmodium* parasites is impacted by STH co-infection in a high transmission setting where the majority of individuals harbor asymptomatic carriage of *P. falciparum*^17^.

Epidemiological studies have estimated the prevalence of STH infection across various regions of Cameroon to be between 0 and 43%, with *Ascaris lumbricoides*, *Trichuris trichiura, Strongyloides stercoralis,* and hookworms being the most prevalent^18, 19, 20, 21, 22, 23^. Co-infection with *Plasmodium falciparum* and STH is common in Cameroon^20, 24, 25, 26, 27^. We have undertaken a longitudinal study of school-aged children in Esse, Cameroon, an area where asymptomatic *P. falciparum* infection is as high as 80%^17^. With this study we specifically sought to evaluate whether STH co-infection and/or exposure may be a driving factor for the asymptomatic malaria phenotype.

## MATERIALS AND METHODS

### Ethics statement

Ethical approvals for this study were obtained from the National Ethics Committee for Human Health Research of Cameroon (1275/CRERSHC/2021 and the University of Utah Institutional Review Board (Protocol number: IRB_00147087). The field studies were also approved by local administrative authorities, including the senior divisional officer in Esse and chiefs of the corresponding villages. Written consent was obtained from participants parents or legal guardians. Cameroonian study personnel who were fluent in the local language and knowledgeable of local customs worked with primary school teachers for the participant consent process and participant sample collection.

### Study Area

This study took place in the Esse Health District in the Centre Region of Cameroon, where malaria transmission is perennial and STHs are regionally endemic^28, 29^. Esse is located at 4°5’38″ N, 11°53’3” E, positioned approximately 657 meters (2155 ft) above sea level, and distanced 83 km NE of the country’s capital city, Yaoundé. Esse Health District is a forest zone with an equatorial tropical climate including two rainy seasons (March-June and September-November) and two dry seasons (July-August and December-February). As of 2005, the estimated total population of Esse was 16,822 inhabitants^30^. The population is generally rural and the predominant productive activity is agricultural.

### Study population

The study population comprised of 134 children, aged 5-15 years, from three primary schools located in Cameroon’s Esse Health District. The three schools included were Ecole publique d’Afanetouana (AF), Ecole publique Ngondi-bele (NG), and Ecole publique d’Etoutoua (ET).

### Study design

This was a 1-month longitudinal study performed from November to December 2021 in Esse Health District (Centre Region, Cameroon) across the three aforementioned primary schools. Demographic measures such as age, biological sex, weight, height, nutritional status, and bed net usage were recorded. An initial temperature check and venous blood samples were collected from all 134 participants and stool samples were collected from all participants able to provide a sample (128 out of 134). Initial data measurements and samples were labeled as occurring at T0. Participants determined to have symptomatic *Plasmodium* infection and/or active intestinal helminthic infection at T0 were notified and treated using the local standard of care practice. Participants found with asymptomatic *Plasmodium* infection at T0 were followed over the course of a month, with daily temperature checks performed by their school teacher to closely evaluate transition from asymptomatic to symptomatic *Plasmodium* infection. For any child who developed symptomatic *Plasmodium* infection during the study period, the child was designated as having temporary asymptomatic malaria. Repeat venous blood samples were collected within 24 hours of developing a fever (Tc) and treatment according to local standard of care practices was given. At the end of the 1-month longitudinal study, all children determined to have malaria, if not already treated, were given anti-malarial treatment according to local standard of care practices regardless of symptoms. Endemic STH species were confirmed by the local public health department. Indirect ELISA methods measuring total IgG for *Trichuris, Ascaris,* and *Strongyloides* were performed using the archived plasma specimens from all 134 children collected at T0 and, if applicable, at Tc.

### Definitions

In accordance with local medical-care settings, a fever was defined as a measured axillary temperature of 37.5°C or greater. Asymptomatic *Plasmodium* infection was defined as any participant with evidence of microscopic *Plasmodium* infection without fever or other overt clinical symptoms. Uncomplicated symptomatic *Plasmodium* infection was defined as the presence of fever and identifiable microscopic *Plasmodium* infection without severe clinical manifestations. Participants initially classified as having asymptomatic *Plasmodium* infection who subsequently developed fever during the course of the longitudinal study were designated as having temporary asymptomatic infection. Those with asymptomatic *Plasmodium* infection who did not develop fever or other symptoms during the follow-up period were classified as having persistent asymptomatic infection. Active intestinal helminth infection was defined as any participant with a microscopy stool Ova and Parasite (O&P) test positive for larvae, eggs, and/or adult worms of one or more of the STH of interest (i.e. *Trichuris trichiura, Strongyloides stercoralis, Ascaris lumbricoides).* STH exposure is defined as a child with total IgG serological positivity to the corresponding STH, with or without a positive stool O&P.

### Sample collection and processing

From each participant, approximately 6 mL of venous whole blood was collected in an EDTA-coated tube at T0 and, if applicable, at Tc. When able, a single stool sample was also collected at T0; repeat stool samples were not collected. All collected samples were transported the same day in refrigerated boxes to the Molecular Parasitology Laboratory at Centre Pasteur du Cameroun in Yaoundé, Cameroon. Fresh whole blood samples were immediately used for *Plasmodium falciparum*/pan antigen rapid diagnostic testing (Standard Q Rapid Test, SD Biosensor, Suwon-si, Gyeonggi-do, Republic of Korea) and for blood hemoglobin quantification using an automated hemoglobinometer (Mission Hb, San Diego, CA). In addition, thin and thick blood smears were prepared and analyzed by light microscopy for *Plasmodium* species detection and enumeration. A fraction of each whole blood sample was centrifuged to separate plasma for serological assays. Plasma, along with the remaining whole blood and red cell pellets, were stored at -80° C. Stool samples were analyzed using the Kato Katz method to detect STH infection using standard protocols. Participant anemia severity was determined using the WHO anemia classification scoring^31^ and annotated using 0 = No anemia; 1 = Mild anemia; 2 = Moderate anemia; 3 = Severe anemia.

### Microscopy

Thick and thin blood films were prepared using 5 μl of whole blood and stained with a 10% Giemsa solution and examined under a x100 oil immersion lens. A blood slide was declared positive when a concordant result was obtained by at least two independent microscopists. Slides were declared negative if no parasites were detected after a count of 500 white blood cells (WBC). In accordance with WHO Microscopy Standard Operating Procedure MM-SOP09, parasite density was determined based on the number of parasites per at least 200 WBC count on a thick film, assuming a total white blood cell count of 8,000 cells/mL of whole blood.

### ELISAs

Plasma samples were used to quantify the levels of total IgG reactive to recombinant antigens from *Ascaris*, *Trichuris*, and *Strongyloides*. For *Trichuris* and *Ascaris*, two recombinant *Trichuris muris* (r*Tm*16, r*Tm*WAP) antigens and two recombinant *Ascaris suum (*r*As*37, r*As*24) antigens, which have been previously described as suitable correlates for identifying *T. trichiura* and *A. lumbricoides* exposure in humans, were used.^32, 33, 34, 35^.

Briefly, for *Ascaris and Trichuris* ELISAs, 96-well Polysorp NUNC ELISA plates (Thermo Scientific, Rochester, New York) were first coated with recombinant helminth antigen (0.25 ug/well)^32, 35^. After blocking with 5% non-fat dry milk powder in 0.1 M carbonate buffer (Sigma Aldrich) wells were washed and incubated with 100 µL of diluted plasma for 2 hours at room temperature. Plasma was diluted 1:100. Bound antibodies were detected with an HRP-labeled goat anti-human (IgG Fc) secondary antibody (Invitrogen, Waltham, Massachusetts) and Super Aqua Blue substrate (Invitrogen, Waltham, Massachusetts). Optical Density (OD) values were obtained at 405 nm absorbency per manufacturer specifications. For each recombinant antigen, duplicate plates using identical plate plans were run. Positive IgG cut-off values were calculated per plate using the formula:

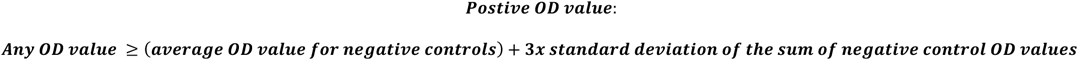

Analysis of *Strongyloides* IgG was performed using a commercially available kit according to the manufacturer’s instructions (GSD01-3012; Gold Standard Diagnostics). *Strongyloides* ELISA results were interpreted according to manufacturer specifications, including Optical Density (OD) values obtained at 450nm-620nm absorbency.

For each antigen, positivity of each sample from the primary plate and the corresponding duplicate plate were compared. Samples which were positive on both plates were labeled as “POSITIVE.” Samples which were negative on both the primary and duplicate plate were labeled as “NEGATIVE.” A total of 39 samples which were discordant between plates were removed from further analysis (13 samples for *As37*, 12 samples for *Tm16*, 11 samples for *Strongyloides*, 1 sample for all 3 antigens, 1 sample for *Tm*16 and *Strongyloides,* and 1 sample for *As*37 and *Tm*16).

OD values were averaged across the primary and corresponding duplicate plate for all confirmed “POSITIVE” and “NEGATIVE” samples for final analysis. Notably, recombinant antigens *As*24 and *Tm*WAP were found to be subsets of *As*37 and *Tm*16, respectively, meaning that all samples positive for *As*24 were also positive for *As*37 and all samples positive for *Tm*WAP were positive for *Tm*16. The reverse was not true (i.e. not all positive *Tm*16 samples were also *Tm*WAP positive and not all positive *As*37 samples were also *As*24 positive). Thus, we proceeded to analyze seroreactivity to *As*37 and *Tm*16.

Of children with temporary asymptomatic malaria, ELISAs for paired samples collected at T0 and Tc were performed with all T0 and Tc samples plated on the same plate for equal comparison. OD values were averaged across duplicate wells prior to calculating OD cut-off values. The change in positivity status

(e.g. positive to negative, negative to positive, etc.) was also recorded for each sample.

### Statistical analysis

Due to the non-normal nature of the data using a Shaprio-Wilk test, data was normalized using log transformation. Furthermore, due to the multivariate nature of the associated measures, linear mixed models were run using the lme4^36^ and lmerTest (3.1-3) packages in R statistical programming language (4.2.2). The response variable was helminth OD value with a fixed effect of malaria status (asymptomatic or symptomatic). Random effects of age, study site, sex, and BMI for age z-score were included in the model due to the unknown variation controlling each. A single model was run for OD value for each soil transmitted helminth (*Trichuris*, *Ascaris* or *Strongyloides*). Due to the lack of variation in the random effects and their control on the outcome of the models, we ran linear models to understand the relationship between helminth OD and parasite density. Data were also compared for significance using paired and non-paired Mann Whitney-U tests or Kruskal Wallis tests in Graph Pad Prism 10.1.0 software (GraphPad Software, LLC, San Diego, CA). Data were plotted using Graph Pad Prism 10.1.0 software (GraphPad Software, LLC, San Diego, CA).

## RESULTS

### Sociodemographic and clinical characteristics of the study population

Of the 134 children included in the study, the majority were male (N = 75, 56.0%), the average age was 10.4 years (range 5 – 15 years), and the average BMI for age Z-score was -0.33. Amongst the three study sites, AF had the most participants (49.3%), followed by NG (34.3%) and then ET (16.4%). Complete demographic data is described in **Table 1**.

**Table 1.**
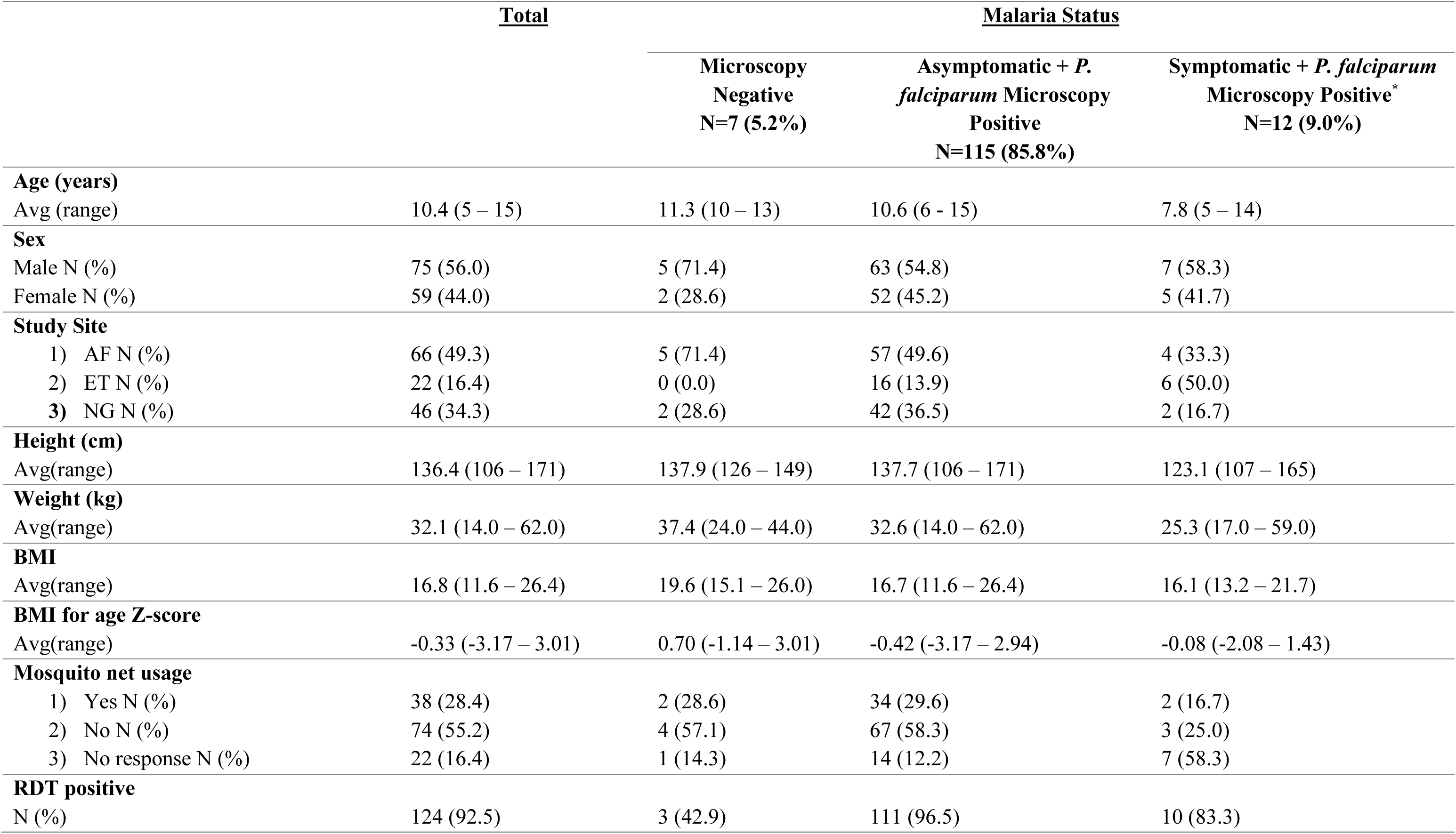
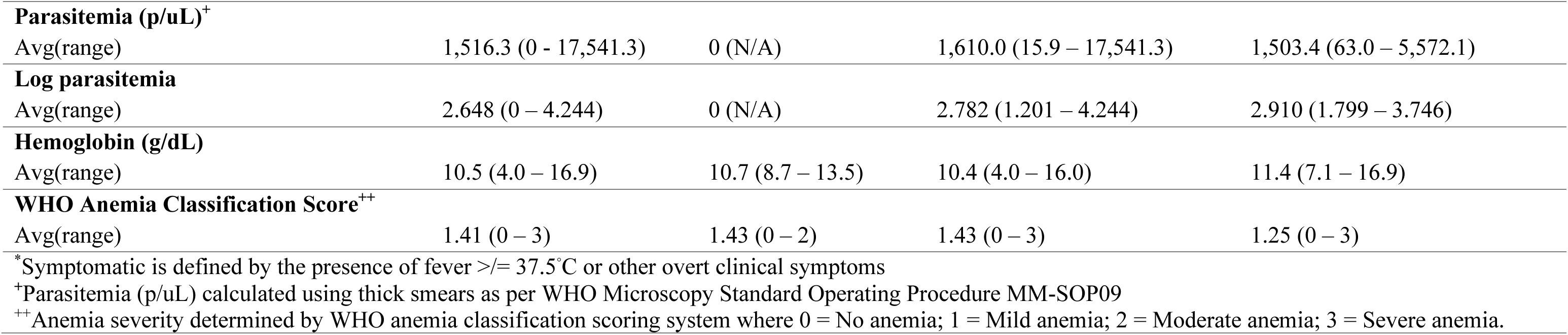
Characteristics of the study population with respect to malaria status (N =134)

### Prevalence of asymptomatic *P. falciparum* infection

Out of the 134 children enrolled in our study, 115 (85.8%) had asymptomatic *P. falciparum* infection by microscopy at the initial time point T0. Of the remaining children, 12 (9.0%) had uncomplicated symptomatic *P. falciparum* infection by microscopy and 7 (5.2%) were microscopy negative for *P. falciparum* (**Fig. 1A**). Of the 127 children with *P. falciparum* infection detected by microscopy, there was no significant difference in the levels of circulating iRBCs in children who presented with asymptomatic malaria compared with those who had uncomplicated malaria (p=0.4677, **Fig. 1B**).

**Figure 1:**
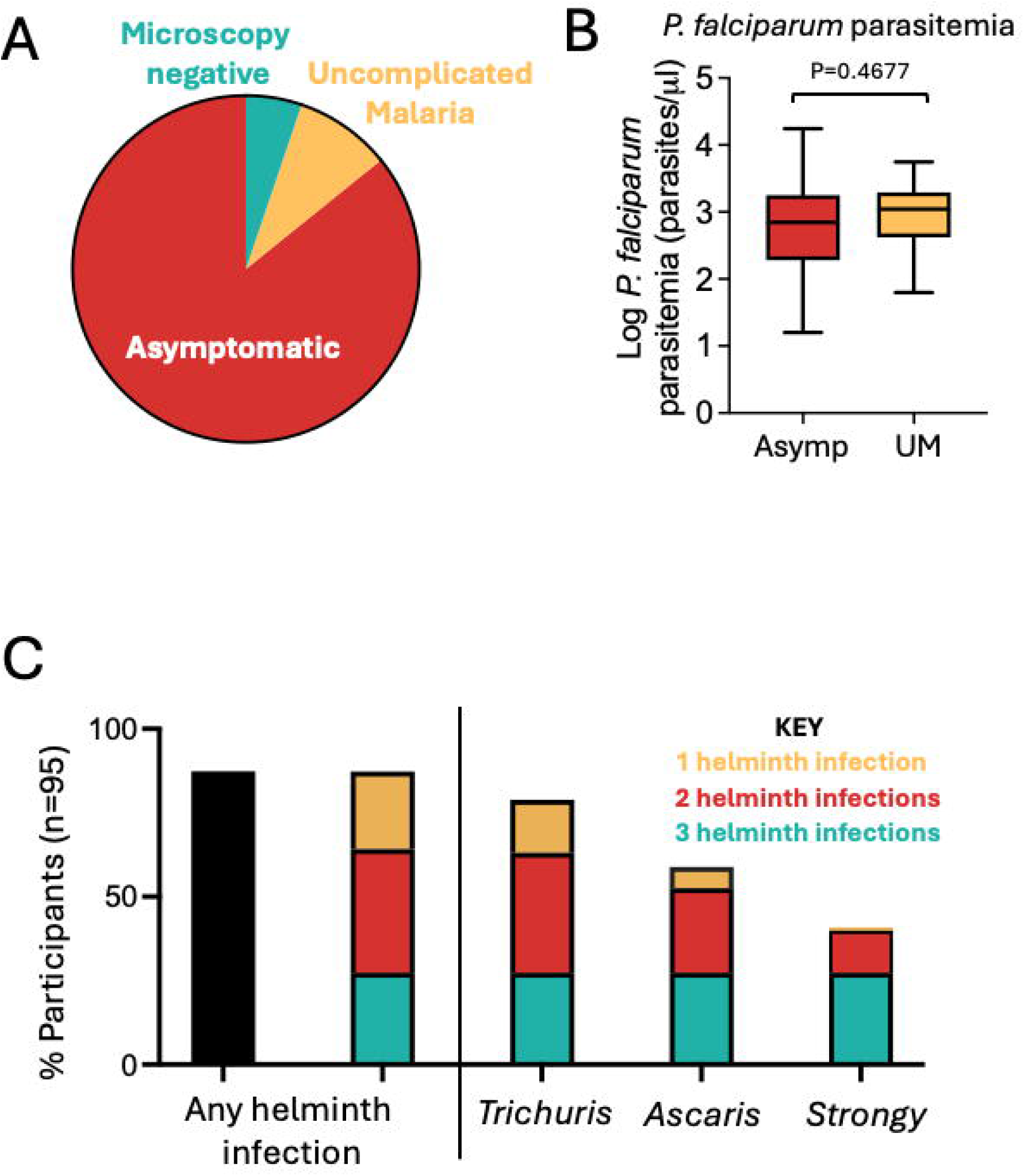
Asymptomatic malaria and STH prevalence are both high in Esse, Cameroon. Prevalence of asymptomatic *P. falciparum* infection (**A**) with associated circulating parasitemia levels (**B**) and the prevalence of STH infection (**C**) in children living in Esse, Cameroon. Bar graph shows the split across all 95 participants with unequivocal ELISA results who were STH positive (black bar) into seropositivity for 1 (gold), 2 (red) or all 3 (turquoise) STH infections. For each individual STH, the red bars represent co-infection with two STH species as follows: *<u>Trichuris</u>*: 24.21% had *Trichuris* and *Ascaris* and 11.58% had *Trichuris* and *Strongyloides*; *<u>Ascari</u>*<u>s</u>: 24.21% had *Trichuris* and *Ascaris* and 1.05% had *Ascaris* and *Strongyloides; <u>Strongyloide</u>s*: 11.58% had *Strongyloides* and *Trichuris* and 1.05% *Strongyloides* and *Ascaris.* In B, parasitemia levels were compared by a Mann Whitney-U test. Asymp: asymptomatic malaria; UM: Uncomplicated Malaria; *Strongy*: *Strongyloides*. <u>Alt text:</u> Panel A shows a pie chart of the proportion of participants who had asymptomatic (85.8%) or uncomplicated malaria (9.00%) or who were microscopy negative for *P. falciparum* (5.2%). Panel B shows a box plot comparing the circulating *P. falciparum* parasitemia in children with asymptomatic or uncomplicated malaria. There was no statistical difference as tested by a Mann Whitney-U test. Panel C shows a compound bar graph that subdivides the 87.4% of participants with seropositivity to soil transmitted helminth infection by the number of co-infecting soil transmitted helminth infections. 76.8% were seropositive for *Trichuris* infection, 52.6% were seropositive for *Ascaris* infection and 37.3% were positive for *Strongyloides* infection. The proportion of individuals with seropositivity to each of the soil transmitted helminth infections is broken down into positivity to all three soil transmitted infections (27.34%) in turquoise, 2 infections (red) or the single infection (gold).

### Prevalence of STH infection and serological positivity

Of the 128 children who provided stools samples, only 3 were positive for STH using Kato Katz (1 *Trichuris*, 2 *Ascaris*). Serologic positivity for at least one STH could not be determined in 39 participants due to equivocal ELISA results for one or more antigens. Of the remaining 95 children, 83/95 (87.4%) had serologic positivity for one or more STH where 22/95 (23.2%) were serologically positive for 1 STH, 35/95 (36.8%) were serologically positive for 2 STH, and 26/95 (27.4%) were serologically positive for 3 STH **(Table 2**, **Fig. 1C**). All 3 children with a positive Kato Katz also had a positive total IgG for the same STH. In this group of 95 children (**Fig. 1C**), the most common STH exposure was to *Trichuris*, detected in 78.9% of children. This included 15.8% who were positive for *Trichuris* alone and 63.2% who were positive for *Trichuris* in combination with *Ascaris* and/or *Strongyloides*. *Ascaris* exposure was detected in 59.0% of children, with 6.3% positive for *Ascaris* alone and 52.6% positive for *Ascaris* together with *Trichuris* and/or *Strongyloides*. *Strongyloides* exposure was detected in 41.1% of children. Of these, 1.1% were positive for *Strongyloides* alone, while 40.0% were positive for *Strongyloides* in combination with *Trichuris* and/or *Ascaris*.

**Table 2.**
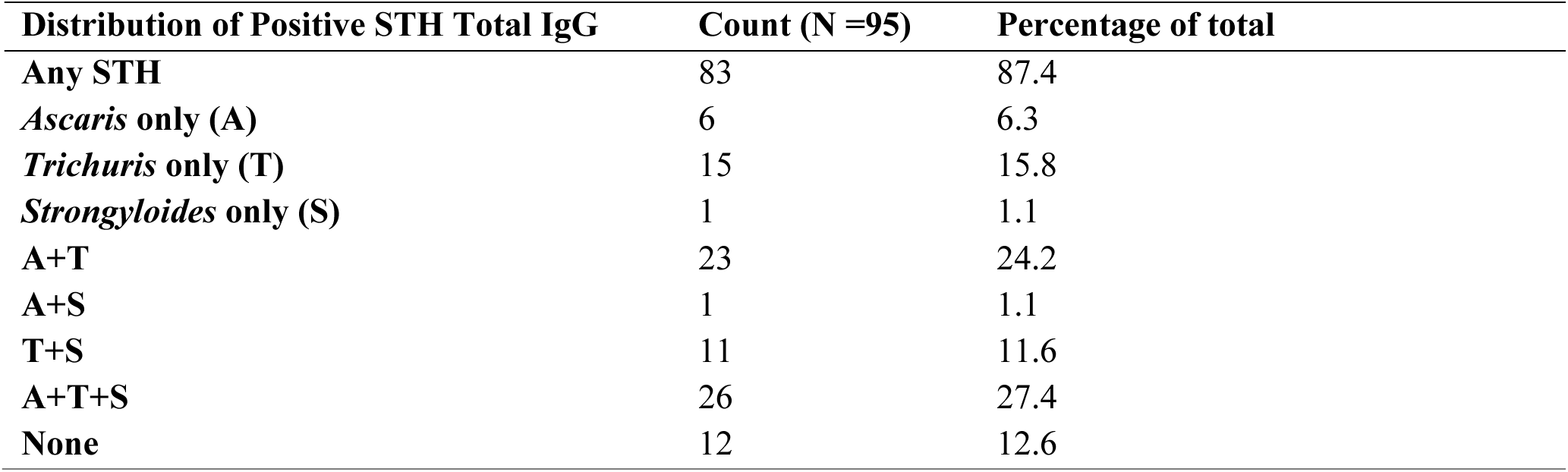
Serologic prevalence of STH using total IgG in children in Esse, Cameroon.

### There was no correlation between serological presence of STH and symptoms of malaria

Under the assumption that STH infection provides protection against the development of symptoms in *Plasmodium* infected children, children with active STH would be expected to be more likely to have asymptomatic malaria relative to those presenting with uncomplicated malaria. Acknowledging the small sample size of children presenting with uncomplicated malaria at the initial time point (N=11 from the 95 children with coherent ELISA results for all three STH infections) and the use of seropositivity here to detect active and past infection, the percentage of children with IgG serological positivity to one or more STH was similar across malarial status (e.g. symptomatic, asymptomatic, microscopy negative) (**Fig. 2A**). Furthermore, the optical density (OD) values for each STH antigen did not significantly differ based on malaria status (Linear Mixed Modelling; p = 0.616 for *Trichuris*, p = 0.749 for *Ascaris*, p = 0.328 for *Strongyloides*) (**Fig. 2B**) suggesting that a higher seroreactivity to STH (and possibly degree of STH exposure) was not protective against development of malarial symptoms. There was also no correlation between the level of seroreactivity to STH and the degree of *P. falciparum* parasitemia for each of the 3 STH studied (Linear Modelling; R^2^ <0.01 p = 0.622 for *Trichuris*, R^2^ <0.01 p = 0.276 for *Ascaris*, R^2^ = 0.01 p = 0.202 for *Strongyloides*) (**Fig. 2C**) suggesting that higher levels of exposure to STH did not influence the ability of *Plasmodium* iRBCs to circulate in the bloodstream in this study.

**Figure 2:**
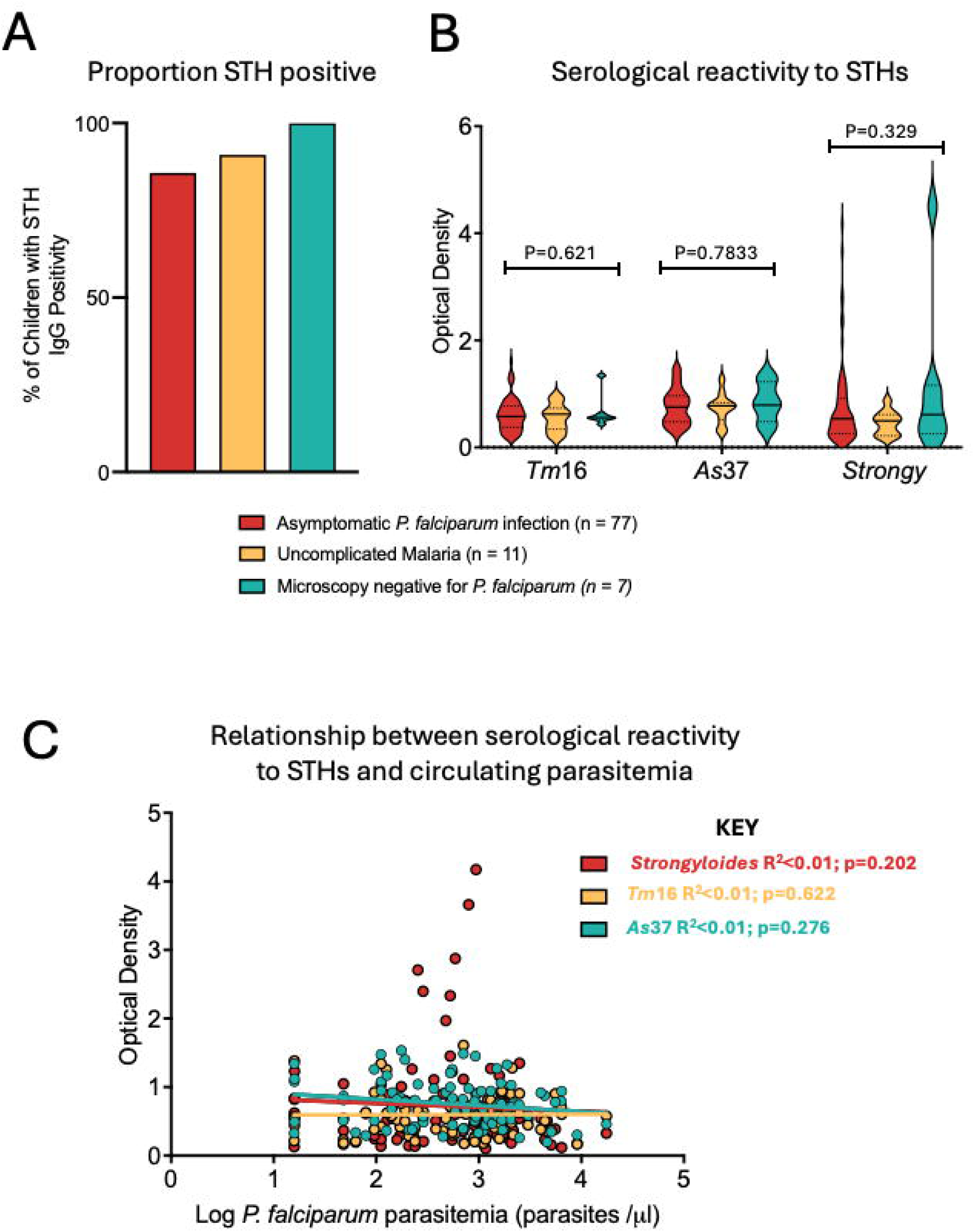
There is no relationship between seropositivity to STH and the presentation of *P. falciparum* malaria. Of children that were seropositive to any STH, the proportion of children with asymptomatic malaria was similar to those with uncomplicated malaria, with 100% of the 7 children in our study who were microscopy negative for circulating *P. falciparum* iRBCs seropositive for STH (**A**). There were no significant differences between the level of seroreactivity to any of the STH with respect to malaria status (**B**). There was also no correlation with the levels of circulating *P. falciparum* iRBCs and the level of seroreactivity to any of the STH in the 88 children who had detectable *Plasmodium* infection (**C**). OD values for each helminth were compared between malaria groups using linear modelling and a Kruskal Wallis multi-comparison test. Correlations between OD values and circulating iRBCs was tested using linear modelling for each STH antigen. *Tm*16: recombinant *Trichuris muris* 16kDa antigen; *As*37: recombinant *Ascaris suum* 37kDa; *Strongy*: recombinant *Strongyloides* antigen. <u>Alt text:</u> Panel A is a bar graph showing that 85.7.4% of children with asymptomatic malaria, 90.9% of children with uncomplicated malaria and 100% of children who were microscopy negative for *P. falciparum* had seropositivity to one or more of the soil transmitted helminths tested. Panel B is a violin plot showing the IgG optical density values for *Trichuris Tm*16 recombinant protein, *Ascaris As*37 recombinant protein and *Strongyloide*s in study participants. No differences for any of the responses were noted for seropositivity to any of the helminth infections with respect to malaria status. Panel C is a scatter plot containing data from participants who were microscopy positive for *P. falciparum* with the IgG optical density values for each of the soil transmitted helminths plotted on the y axis against the density of *P. falciparum* infected red blood cells in the circulation on the x axis. All best fit lines are flat and no significant correlations were found.

### Breadth of exposure to STH species was not correlated with malaria status

Amongst the 95 children with defined exposure to the three STH tested in this study, there was no demonstrable difference between the proportion of children with asymptomatic *P. falciparum* infection who were exposed to STH compared to those who were not exposed to STH (**Fig. 3A**). Although the number of participants with no seropositivity to STHs was small (N=12) this data suggests that asymptomatic malaria can occur independently of STH infection. Given that there was a differential positivity to the number of STH antigens among the participants, we then tested whether asymptomatic malaria might be associated with exposure to a higher number of STH species. However, we did not see any correlation between the breadth of exposure to STH and malaria status (Linear Modelling; R^2^ < 0.01 p = 0.701) (**Fig. 3B**) suggesting that exposure to several different species of STH is also not a necessary pre-requisite to development asymptomatic malaria.

**Figure 3:**
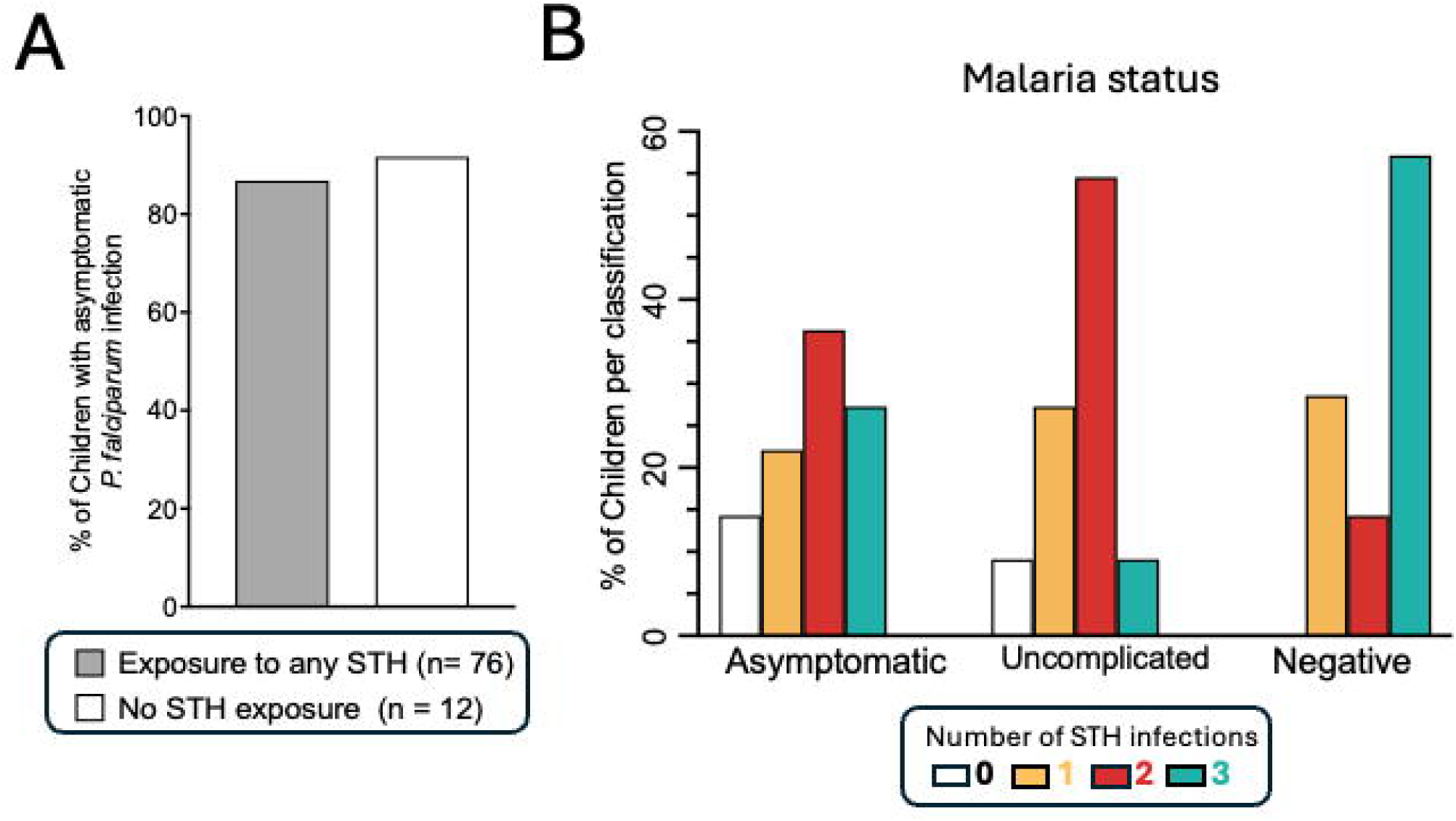
Serological evidence of STH exposure does not correlate with symptoms of malaria. The proportion of children with asymptomatic malaria is similar between those who are STH positive (86.8%) and those who are STH negative (91.7%) (**A**). The number of species of STH exposure does not correlate malaria phenotype (**B**). The correlation of breadth of exposure to STH with symptoms of malaria was performed with linear modelling. <u>Alt text:</u> Panel A is a bar graph showing that 86.4% of asymptomatic children had serological exposure to soil transmitted helminths and 13.6% of asymptomatic children were serologically negative for soil transmitted helminths. Panel B is a bar graph which breaks down the number of soil transmitted helminths that children were seropositive for by malaria status. There was no association between the number of soil transmitted helminth infection exposures and the malaria status of the participants. 14.3% of children with asymptomatic malaria had no soil transmitted helminth infections and 22.1% were positive for 1 infection, 36.4% were positive for 2 infections and 27.3% were positive for all three soil transmitted helminth infections. 9.1% of children with uncomplicated malaria had no serological evidence for soil transmitted helminth infections with 27.3% seropositive for 1 infection, 54.5% seropositive for 2 infections and 9.1% seropositive for all three soil transmitted helminth infections. None of the children negative for *P. falciparum* blood stage infection had soil transmitted infections, with 28.6% seropositive for 1 infection, 14.3% seropositive for 2 infections and 57.1% seropositive for all three soil transmitted helminth infections.

### There was no correlation between STH seropositivity and the development of malaria symptoms in children with temporary asymptomatic malaria

Of the 115 children determined to have asymptomatic *P. falciparum* infection from the cohort of 134 children, 14 (12.2%) developed a fever >/= 37.5^◦^C during the study. These participants were labeled as having temporary asymptomatic *Plasmodium* infection meaning that they converted from having asymptomatic *P. falciparum* infection to uncomplicated symptomatic *P. falciparum* infection during the study timeline. Demographics for children with temporary asymptomatic infection are detailed in **Table 3**. The average number of days to development of symptoms was 8 days (range 3-17 days). One of the 14 children with temporary asymptomatic infection was too ill at the time of conversion to further participate and was thus removed from the study at that point and directed to appropriate medical care. Of the remaining 13 children with temporary asymptomatic *Plasmodium* infection, none had evidence of active STH infection on initial Kato Katz stool testing. At the point of temperature development (Tc), 9 children were serologically positive for *Strongyloides*, 2 were positive for *Trichuris* and 2 were positive for *Ascaris* (**Fig. 4**). STH IgG OD values from T0 were compared to IgG OD values from Tc and were significantly higher for *Tm*16 (p =0.0005) (**Fig. 4A**), *As*37 (p=0.0002) (**Fig 4B**), and trending towards increased for *Strongyloides* (p = 0.0681) (**Fig. 4C**). This data does not support the hypothesis that symptoms of malaria may be associated with lack of a STH infection.

**Figure 4:**
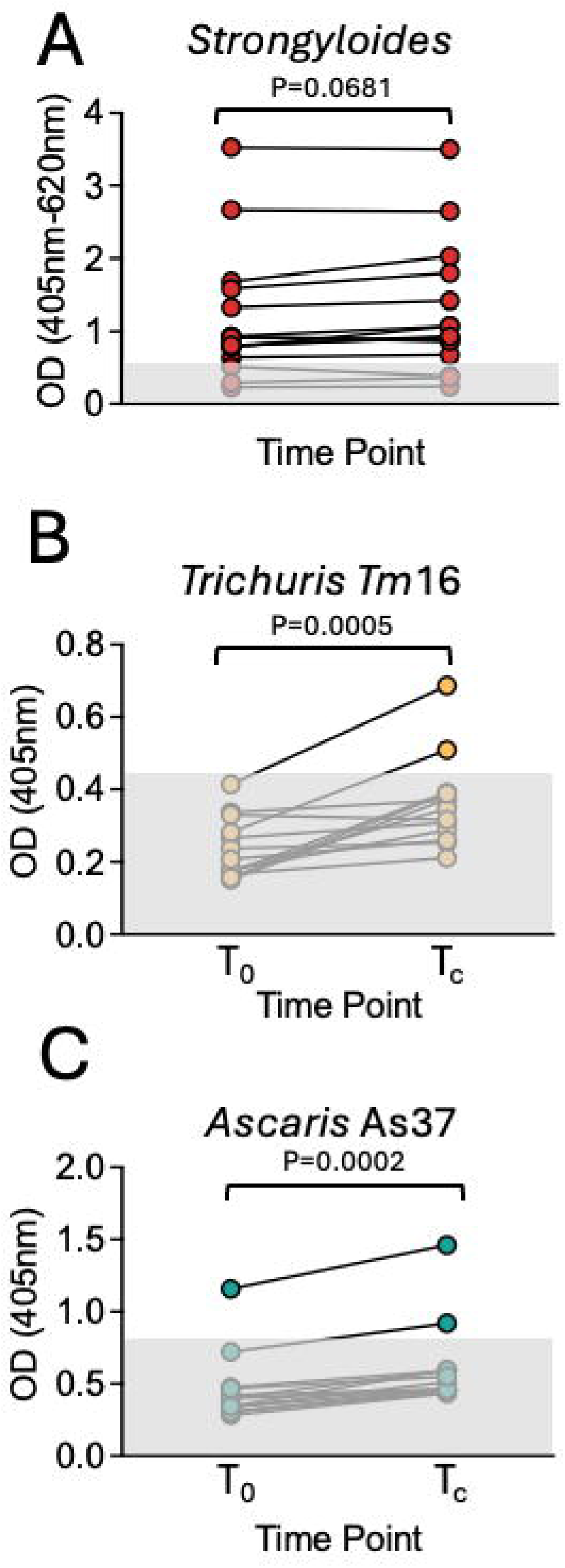
The serological levels of antibody to recombinant STH antigens in asymptomatic children does not decrease upon development of uncomplicated malaria. Paired samples of children from the initial time point of the study (T0) and within 24 hours of developing a fever (Tc) over the course of the 1-month long study were tested for change in serological responsiveness to *Strongyloides* (**A**), *Trichuris muris* recombinant *Tm*16 (**B**) and *Ascaris suum* recombinant *A*s37 (**C**). The shaded box shows the range of negative OD values that were σ; (average OD value for negative controls +3x standard deviation of the sum of negative control OD values). Wilcoxon matched pairs signed rank tests were used to compare differences before and after developing uncomplicated malaria. <u>Alt text:</u> Paired x-y plots with connecting line for each child show for the optical density of IgG responses to *Strongyloides* (top), *Trichuris* (middle) and *Ascaris* (bottom) before and after developing a fever. The data shows that once children develop malarial fever, seropositivity either remains the same (*Strongyloides*; p=0.0681) or increases (*Trichuris* and *Ascaris*; P<0.0005). Statistical significance was tested using Wilcoxon matched pairs signed rank tests.

**Table 3.**
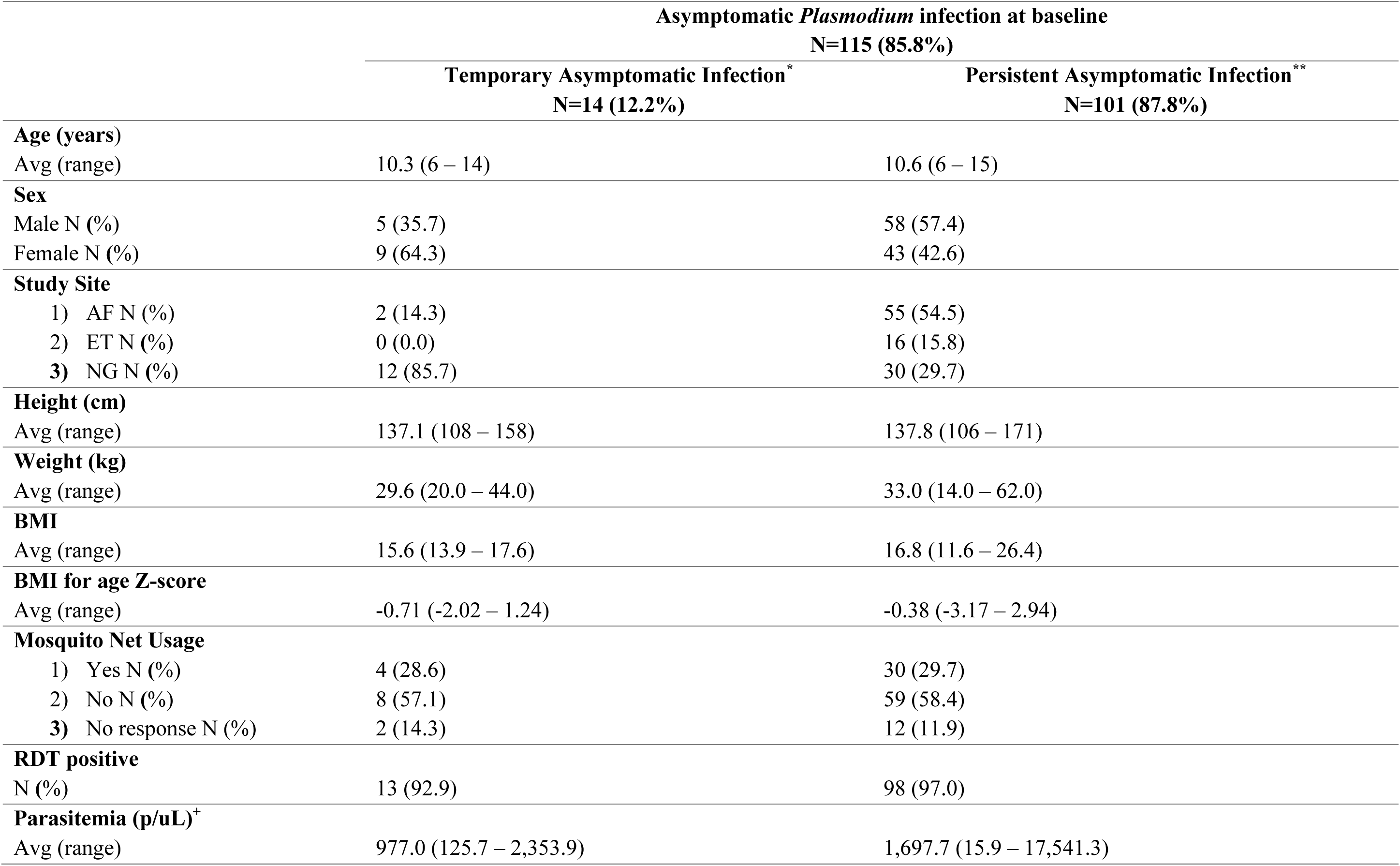
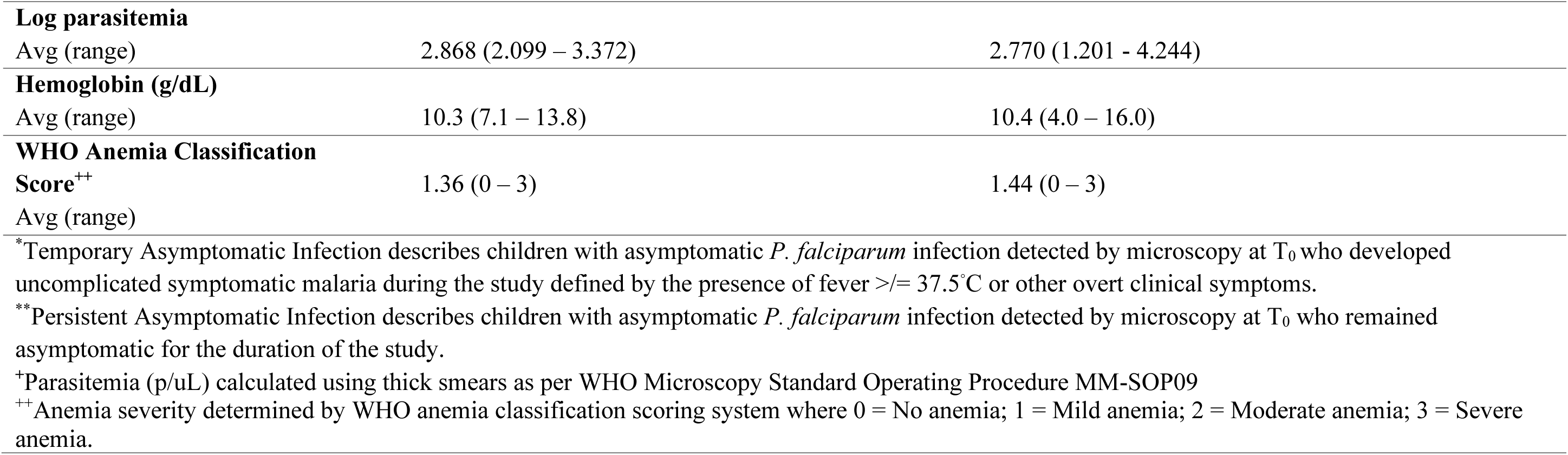
Risk factors associated with development of symptoms in participants with asymptomatic P. falciparum infection.

|  | Asymptomatic *Plasmodium* infection at baseline |  |
| --- | --- | --- |
|  | N=115 (85.8%) |  |
|  | Temporary Asymptomatic Infection* | Persistent Asymptomatic Infection** |
|  | N=14 (12.2%) | N=101 (87.8%) |
| **Age (years)** |  |  |
| Avg (range) | 10.3 (6 – 14) | 10.6 (6 – 15) |
| **Sex** |  |  |
| Male N (%) | 5 (35.7) | 58 (57.4) |
| Female N (%) | 9 (64.3) | 43 (42.6) |
| **Study Site** |  |  |
| 1) AF N (%) | 2 (14.3) | 55 (54.5) |
| 2) ET N (%) | 0 (0.0) | 16 (15.8) |
| 3) NG N (%) | 12 (85.7) | 30 (29.7) |
| **Height (cm)** |  |  |
| Avg (range) | 137.1 (108 – 158) | 137.8 (106 – 171) |
| **Weight (kg)** |  |  |
| Avg (range) | 29.6 (20.0 – 44.0) | 33.0 (14.0 – 62.0) |
| **BMI** |  |  |
| Avg (range) | 15.6 (13.9 – 17.6) | 16.8 (11.6 – 26.4) |
| **BMI for age Z-score** |  |  |
| Avg (range) | -0.71 (-2.02 – 1.24) | -0.38 (-3.17 – 2.94) |
| **Mosquito Net Usage** |  |  |
| 1) Yes N (%) | 4 (28.6) | 30 (29.7) |
| 2) No N (%) | 8 (57.1) | 59 (58.4) |
| 3) No response N (%) | 2 (14.3) | 12 (11.9) |
| **RDT positive** |  |  |
| N (%) | 13 (92.9) | 98 (97.0) |
| **Parasitemia (p/uL)<sup>+</sup>** |  |  |
| Avg (range) | 977.0 (125.7 – 2,353.9) | 1,697.7 (15.9 – 17,541.3) |
| **Log parasitemia** |  |  |
| Avg (range) | 2.868 (2.099 – 3.372) | 2.770 (1.201 - 4.244) |
| **Hemoglobin (g/dL)** |  |  |
| Avg (range) | 10.3 (7.1 – 13.8) | 10.4 (4.0 – 16.0) |
| **WHO Anemia Classification** |  |  |
| **Score<sup>++</sup>** | 1.36 (0 – 3) | 1.44 (0 – 3) |
| Avg (range) |  |  |
| *Temporary Asymptomatic Infection describes children with asymptomatic *P. falciparum* infection detected by microscopy at T<sub>0</sub> who developed uncomplicated symptomatic malaria during the study defined by the presence of fever $\geq 37.5^{\circ}\text{C}$ or other overt clinical symptoms. | | |
| **Persistent Asymptomatic Infection describes children with asymptomatic *P. falciparum* infection detected by microscopy at T<sub>0</sub> who remained asymptomatic for the duration of the study. |  |  |
| <sup>+</sup>Parasitemia (p/uL) calculated using thick smears as per WHO Microscopy Standard Operating Procedure MM-SOP09 |  |  |
| <sup>++</sup>Anemia severity determined by WHO anemia classification scoring system where 0 = No anemia; 1 = Mild anemia; 2 = Moderate anemia; 3 = Severe anemia. |  |  |

## Discussion

STH are a highly prevalent parasitic diseases affecting – and imposing a high burden on - individuals of all ages across many low- and middle-income countries^37, 38, 39^. Many areas with high STH burden are also impacted by the widespread prevalence of asymptomatic *P. falciparum* carriage in adults and children alike ^17, 40, 41, 42, 43, 44^. Whilst individuals over the age of 15-20 years old have been shown to have superior control of *P. falciparum* iRBCs due to the development of robust^45, 46^ and broadly-reactive^47, 48^ humoral responses, the host factors which facilitate asymptomatic carriage of *P. falciparum* infection in children who have not yet developed this level of humoral immunity are not well understood. Given that children often bear the burden of STH infections, we sought to evaluate whether infection with or without exposure to STH infection is associated with asymptomatic*. P. falciparum* infection.

The immunological impact of helminth infections on the host is well described with the majority of studies suggesting an overall anti-inflammatory host environment created via both innate and adaptive mechanisms. This includes the induction of Type 2 and regulatory T cell responses and modulation of macrophages and dendritic cells resulting in higher levels of IL-10 and TGF-B.^26, 49, 50^. The impact of the helminth-induced immune regulation has been found to play a role in vaccination responses^51, 52, 53, 54^, outcomes of co-infections^26, 50^, and modified atopic responses^55, 56^. Cases of helminth co-infection and asymptomatic *Plasmodium* infections in children have been described in endemic regions^24, 25, 57, 58^. Mouse and human studies evaluating the impact of helminth co-infection on *Plasmodium* infection have observed mixed results with regards to clinical outcomes. Depending of species of murine *Plasmodium*, this spans protective effects^26, 50, 59^ through a negligible impact^50^ to increased clinical severity^27, 50^.

Without treatment, helminth infections can be chronic lasting from weeks to years^7, 8, 9^. Children living in endemic areas are particularly affected with prevalence ranging from ∼26% to as high as 67% depending on geographical location^60^. Based on a general understanding of how helminths modulate inflammatory responses to facilitate their survival it has been hypothesized that spill-over of immunoregulation from chronic helminth infections may also dampen harmful inflammatory responses to *P. falciparum* iRBCs, in turn facilitating the asymptomatic carriage of *Plasmodium* parasites. One study in Ghanaian children demonstrated that the cellular immune response to *P. falciparum* antigens in helminth-infected children significantly differed from helminth non-infected children^57, 58^. Specifically, there was elevated IL-10 and increased expression of suppressive cytokine signaling molecules SOCS-3, FOXP3, and PD-1 in peripheral blood mononuclear cells (PBMCs) stimulated with *P. falciparum* iRBCs in the co-infected group compared to the *P. falciparum*-only infected group. This study supported the hypothesis that helminth infection may foster asymptomatic carriage of *P. falciparum* infection, identifying potential mechanisms by which helminth infections could reduce *P. falciparum*-induced inflammation.

Here we have used serological analysis to determine whether STH exposure overlaps with asymptomatic *P. falciparum* infection in a high transmission area of Cameroon. We had very few Kato Katz positive children (2.3%). This is significantly less than expected for an endemic region where local health authorities report a STH prevalence of 28-47%. However, stool Kato Katz is frequently unreliable with a positivity rate of 55% in individuals with three separate specimens submitted for O&P examination to as low 19.8% when only one stool specimen is submitted, even in high prevalence settings^61^. Serological determination of STH exposure, whilst unable to definitively distinguish active infection from past exposure, facilitates a more long-term picture of exposure over time and is more likely to indicate past infection as this area (Esse Health District) has been treated with ivermectin against onchocerciasis since 2011 and it has been clearly demonstrated that ivermectin has a collateral impact on STH^62^. Although in endemic areas the potential for co-infection with multiple STH exists and certain structural similarities between parasites can result in serological cross-reactivity and subsequent false positive results, use of recombinant antigens helps to reduce this issue. We utilized the recombinant *Tm*16^33^ and *As*37^32^ antigens for our analysis and we verified our results with recombinant *Tm*WAP^35^ and *As*24^34, 63^ antigens. The commercially available Gold Standard Diagnostics *Strongyloides* IgG ELISA kit we used also claims the use of recombinant antigen. An additional diagnostic complication when depending on serological positivity is seroreversion (i.e. antibody positive to antibody negative). There are few studies on seroreversion rates in *Trichuris* infection, however it is known that a proportion of individuals with *Ascaris*^64^ and *Strongyloides* ^65, 66, 67^ infections show IgG seroreversion within 6 months of receiving targeted anti-helminthic therapy. This makes determination of active infection, onset of infection, and resolution of infection very difficult to establish but does give a general time frame by which many individuals are likely to have been infected if serologically positive. The lack of symptomatic disease in most children with active STH infection also inhibits the ability to estimate the presence of active infection and its onset.

Our study demonstrates that IgG serologic positivity to STH including *Ascaris*, *Trichuris*, and *Strongyloides* and asymptomatic *P. falciparum* infection are highly prevalent in generally healthy, school-age children residing in Esse, Cameroon. We identified that 69.5% of the 95 children with unequivocal serological tests had both asymptomatic *P. falciparum* infection and STH IgG positivity (**Fig. 2A**). Despite this high rate of co-infection, our study sheds some doubt on the hypothesis that STH exposure is a key component in the facilitation of asymptomatic carriage of *P. falciparum* iRBCs. At our initial time point 91.7% of symptomatic individuals with uncomplicated malaria were also serologically positive for STH (**Fig. 2A**). Furthermore, the breadth of STH infection was similar between asymptomatic and uncomplicated malaria groups (**Fig. 3B**). We also saw that, upon developing uncomplicated malaria, children who were previously asymptomatic had a higher rather than lower serological reactivity to *Trichuris* and *Ascaris* and a propensity towards a higher rather than lower serological reactivity to *Strongyloides* (**Fig. 4**). Additionally, all 13 of the children with temporary asymptomatic malaria had negative stool microscopy testing at the T0, suggesting against an overwhelming STH burden initially, although these could potentially represent false-negative tests given the low sensitivity of stool O&Ps with only a single specimen and our limited resource setting.

Another area of potential ambiguity in our results relates to fever, particularly the measurement of fever as well as the presumed etiology of fever. Fever in our study was defined as an axillary temperature of 37.5^◦^C or greater in accordance with local clinical standards. Per thermometer manufacturer guidelines, 0.5^◦^C was added to the measured body temperature in order to determine the child’s actual body temperature. Multiple children included in our study who were classified as febrile, including both those children with fever at T0 and Tc, had a borderline temperature of 37.5^◦^C. It is possible that the number of truly febrile children in our study is overestimated due to temperature calculation based on manufacturer assumption, as well as the relatively low threshold temperature to define a fever. In the United States the American Academy of Pediatrics defines the threshold for fever in older children, to be 38.3^◦^C^68^. Furthermore, while it is generally agreed in clinical settings that rectal temperature recordings are the gold standard for measuring true or core body temperature and that axillary temperatures are consistently lower than rectal temperatures, there is no standard conversion factor to estimate core temperature due to the wide variation in rectal-axillary temperature difference^69^. In our study we assume, in accordance with local clinical practice, that a fever in the setting of *P. falciparum* infection is due to symptomatic malaria until proven otherwise^70^. However it is possible that the fever measured may be due to an alternative etiology such as the a direct result of new helminth infection, at least for *Ascaris*^69, 71^ or *Strongyloide*s^72^ infections, although such STH infections when symptomatic are often accompanied by gastrointestinal symptoms such as diarrhea and abdominal pain and/or, in the case of the former, respiratory symptoms including cough, wheezing, and shortness of breath. Viral and/or bacterial infections^70, 73, 74, 75^, as well as elevated temperature from heat exhaustion and increased activity level while running on a playground outside at school in a hot, humid climate^76, 77^ are also possible causes of fever.

The hypothesis that STH induce immunoregulatory mechanisms capable of suppressing the inflammatory responses driving symptoms of malaria in *P. falciparum* infection may also be expected to manifest in the suppression of mechanisms of parasite control. For example, production of antibodies via differentiation of plasma cells^78, 79^ or activation of phagocytic macrophages to clear iRBCs^80, 81^ may be suppressed leading to a higher circulating parasitemia. However, we did not see any evidence that there was any correlation between serological positivity to any of the three STH and circulating parasite density in our study (**Fig. 2C**).

Though we cannot draw definitive conclusions regarding direct correlations between asymptomatic carriage of *P. falciparum* infections and active STH infections our data suggests that a causal link between asymptomatic *P. falciparum* infection and STH infection in children is unlikely and certainly is not essential. The limitations and the scope of this study do not allow us to draw definitive conclusions on the specific host immunological factors governing asymptomatic *P. falciparum* infection in children. However, a better understanding of the host immunological mechanisms that facilitate pediatric asymptomatic malaria is necessary to successfully achieve disease eradication as well as to understand the consequences of possible secondary impacts of asymptomatic malaria in terms of vaccine efficacy in malaria-endemic areas.

## Data Availability

All data produced in the present study are available upon reasonable request to the authors

## Acknowledgements

We extend our gratitude to the children and teachers at Ecole publique d’Afanetouana (AF), Ecole publique Ngondi-bele (NG), and Ecole publique d’Etoutoua (ET) for their participation in this study.

## Author contributions

LL performed the ELISAs, analyzed the data and wrote the manuscript. BF, AJ, CNM and CS collected the field samples. DA performed the ELISAs. DHC undertook statistical analysis of the data. CD collected the data on circulating parasite density. HCND helped devise the study, served as STH epidemiology expert for Centre Region of Cameroon, and reviewed the paper; BZ provided the recombinant proteins and reviewed the paper; POKN supervised the study and reviewed the paper; LSA devised the study, supervised the study, analyzed the data and wrote the manuscript; TJL devised the study, supervised the study, analyzed the data and wrote the manuscript.

